# Ranking videolaryngoscopes and direct laryngoscopes by orotracheal intubation performance in children: protocol of a systematic review and network meta-analysis of randomized clinical trials at patient level

**DOI:** 10.1101/2021.01.25.21250439

**Authors:** Clístenes C Carvalho, Stéphanie LPA Regueira, Ana Beatriz S Souza, Lucas MLF Medeiros, Marielle BS Manoel

## Abstract

**Background:** Videolaryngoscopy was shown to improve glottic visualization in children as compared to direct laryngoscopy, but at the expenses of delayed time for intubation. As little evidence is available regarding the relative performance of different laryngoscopes at present, we designed this systematic review and network meta-analysis to rank the different videolaryngoscopes (VLs) and direct laryngoscopes (DLs) for orotracheal intubation in children.

**Methods:** We will conduct a search in PubMed, LILACS, Scielo, Embase, Web of Science, and Cochrane Central Register of Controlled Trials (CENTRAL; 2021, Issue 1) on 27/01/2021. We will include randomized clinical trials fully reported with patients aged ≤ 18 years, making comparisons between different types of laryngoscopes (any of both VLs and DLs) for failed first intubation attempt, intubation time, number of attempts at intubation or number of unsuccessful intubations, failed intubation, glottic view score, or adverse responses to endotracheal intubation. Pooled effects will be estimated by both fixed and random-effects models and presented according to qualitative and quantitative heterogeneity assessment. Sensitivity analyses will be performed as well as a priori subgroup, meta-regression and multiple meta-regression analyses. Additionally, network meta-analyses will be applied to rank the different VLs and DLs. We will also assess the risk of selective publication by funnel plot asymmetry.

**Discussion:** This systematic review and network meta-analysis aim to understand which laryngoscopes perform better than others for orotracheal intubation.

**Systematic review registration:** The current protocol was submitted to PROSPERO on 25/01/2021.

## INTRODUCTION

Endotracheal intubation is an important procedure in children to secure airway path and respiratory function for both elective and urgent situations. The most widely used technique to intubate children is the direct laryngoscopy (DL), mostly because of its broad availability.^1,2^ However, to placing a tube into the trachea by a DL is not always an easy task, mainly for infants and small children – who have anatomical particularities that make their airway manipulation even more challenging.^2,3^ Therefore, as difficult intubations are difficult to anticipate, pediatric anesthetists should always be prepared to tackle such situations.^4,5^

To help handling with difficult intubations by DL, pediatric anesthetists can make use of videolaryngoscopes (VLs). These devices were demonstrated to improve larynx visualization and reduce the incidence of failed intubations, airway trauma, and hoarseness in adults.^6^ However, two recent systematic reviews have caught little to no difference between videolaryngoscopy (VL) and DL for many outcomes in children such as intubation time, number of intubation attempts, and lowest oxygen saturation.^7,8^ Both pediatric reviews, however, were made up of few and heterogeneous studies. They also did not evaluate the repercussion of individual VLs over the assessed outcomes, although research has demonstrated different VLs to have differential performance.^6,9^ This way, network analyses assessing both direct and indirect comparisons between VLs and between VLs and DLs might provide more data for summarization besides enabling to rank the different types of devices.

We then designed this systematic review with network meta-analyses to rank VLs and DLs for failed first intubation attempt, intubation time, number of attempts at intubation or number of unsuccessful intubations, failed intubation, glottic view, and some adverse events.

## METHODS

### Protocol and registration

The current protocol was designed according to recommended standards and reported as per the Preferred Reporting Items for Systematic Reviews and Meta-analysis Protocols (PRISMA-P) guidelines.^10-12^ This review protocol was also submitted to PROSPERO registration on 25/01/2021.

### Eligibility criteria

Inclusion criteria will be as follows: 1) randomized clinical trials fully reported; 2) patients ≤ 18 years – not manikins; 3) data available on failed first intubation attempt; intubation time; number of attempts at intubation or number of unsuccessful intubations; failed intubation; glottic view score; or adverse responses to endotracheal intubation 4) comparison between different types of laryngoscopes (any of both VLs and DLs). Exclusion criteria will be as follows: 1) studies published in language other than English, Spanish or Portuguese; 2) impossibility of abstracting relevant data on outcomes at patient level; 3) differences in the intubation technique between intervention groups other than the laryngoscopes.

### Information sources

We will conduct a computerized search through PubMed, LILACS, Scielo, Embase, Web of Science, and Cochrane Central Register of Controlled Trials (CENTRAL) on 27/01/2021. We will also search the reference lists of included studies.

### Search strategy

The following searching strategy line will be applied to databases with no limitations: *(laryngoscopes[MeSH] OR laryngoscopy[MeSH] OR laryngoscop* OR videolaryngoscop* OR GlideScope OR Pentax OR C-MAC OR blade OR McGrath OR X-lite OR Airtraq OR Trueview OR CEL-100 OR “King vision” OR Bullard OR Venner OR vividtrac OR “copilot VL” OR “ue?scope”) AND (“Airway management”[MeSH] OR “intubation, intratracheal”[MeSH] OR “Airway management” OR intubation* OR difficult* OR visualization OR view) AND (child[MeSH] OR pediatrics[MeSH] OR infant[MeSH] OR child* OR pediatric* OR infant OR newborn* OR neonate*)*

### Study records

Retrieved references will be taken to EPPI Reviewer Web (Beta)^13^ for screening steps – “title and abstract” then “full text”. Eligibility criteria will be applied to select the studies to be included. Two doubles of reviewers will perform in duplicate and independently all steps from screening of title and abstract, through screening of full text and risk of bias assessment to data extraction. The results will be compared and disagreements solved by discussion and consensus amongst correspondent researchers and the first author (CC). In case of no consensus, CC will act as final judge. In case of missing data, corresponding authors will be reached. If relevant data is missing or data are conflicting and corresponding author do not reply our contact after three attempts, then the article will be excluded. Data will be recorded in an excel spreadsheet. The data-extraction form will first be tested in 5 included studies and then refined if necessary.

### Data items

We will collect data on first authors name, publication year, study design, characteristics of population, mean age, mean BMI, mean weight, mean height, sex frequencies, ASA physical status, setting, country, sampling, nature of procedure (elective vs urgent), intubation technique (regular vs rapid sequence induction vs awake), manipulator experience, number of participants randomised and analysed, number of participants in each arm, type of laryngoscope, inducer used and dose, opioid used and dose, neuromuscular blocking agent used and dose. Also, for continuous outcomes, mean and standard deviation in each arm and mean difference and standard error between groups. For categorical outcomes, number of events and number of patients in each arm along with relative risk and standard error between groups.

### Outcomes with prioritization

We will primarily investigate the risk of failed first intubation attempt. To getting additional information about how different laryngoscopes perform during orotracheal intubation, we will also evaluate their effects over intubation time, number of attempts at intubation or number of unsuccessful intubations, failed intubation, glottic view – assessed by a validated score – and presence of adverse events such as oxygen desaturation, airway trauma and changes in heart rate or blood pressure. All outcomes will be considered according to studies’ definitions and the repercussions of different definitions over the results will be assessed by sensitivity analyses.

### Risk of bias in individual studies

We’ll apply the RoB 2 tool to assessing the risk of bias of the individual studies for each outcome.^14^ Five domains are assessed through this tool: randomization process; deviation from intended interventions; missing outcome data; measurement of the outcome; and selection of reported results. We will also use risk of bias judgments for sensitivity analysis.

### Data synthesis

Data will be summarized if at least two different sources are available. Analyses will be conducted in Review Manager^15^ (RevMan, London, UK, v5.3.5) and R software tools^16^ (R Foundation for Statistical Computing, Vienna, Austria), as appropriate. Per-protocol data on patient level will be extracted or calculated from studies and used for summarizations. Effect sizes, SE, and 95% CI will be estimated for each study from the recorded data. Forest plots of relative risk or mean difference will be constructed for every outcome. Pooled estimates will be calculated by both fixed-effects (Mantel-Haenszel or inverse variance method, where appropriate) and random-effects (Sidik-Jonkman method with Hartung-Knapp adjustment) for sensitivity analyses. Heterogeneity will be evaluated qualitatively and quantitatively by Cochran’s Q-test, *I*^*2*^, and Graphic Display of Heterogeneity (GOSH). Where qualitative or quantitative heterogeneity is present, pooled estimates from random-effects models will be presented. An influence analysis by Leave-One-Out method will be performed to assess the influence of each study in the pooled effects and the between studies heterogeneity. GOSH plots will also be used to search for subclusters of different effects sizes in order to support the subgroup evaluations. Sensitivity analyses will be undertaken throughout both subgroup and meta-regression analyses. Subgroup assessments will be performed using either mixed-effects or random-effects models, where appropriate, for outcomes with 10 or more studies available. Meta-regressions of single features, one at a time, as well as multiple meta-regression with maximum likelihood (ML) estimator for *τ*^2^ will be conducted only for outcomes with 10 or more studies available per covariate. The multiple meta-regression models will be submitted to a permutation test to confirm statistical significance. Both subgroup and meta-regression analyses will be performed with a priori hypotheses – attempting to avert the catch of spurious associations – with the following features: patients’ age, manipulator experience, intubation technique, population characteristics, setting, nature of procedure, type of laryngoscope, inducer, opioid, and neuromuscular blocking agent used. Risk of bias judgments, different outcome definitions, and decisions made throughout the statistical analysis will also be included in sensitivity analyses. Random-effects network meta-analyses among the different VLs and DLs will also be performed. Network models’ consistency will be tested by net heat plots and net splitting method. Additionally, to deal with the risk of type-I and type-II errors due to repeated significance testing by subsequent meta-analyses, we will apply the trail sequential analysis for the main outcome.

### Meta-bias

We will perform assessment of selective publication by small sample bias methods for those outcomes with 10 or more studies. Funnel plots will be built up and Egger’s tests performed to check for plot asymmetry. The threshold of significance will be set at p<0.100 for this method as this test has low power. Where asymmetries are present, a Duval and Tweedie’s trim-and-fill procedure will be applied to estimate bias-corrected effects.

### Confidence in cumulative evidence

To assess the quality of the evidences, we will apply the Grading of Recommendations, Assessment, Development and Evaluation (GRADE) approach. This approach takes into account factors related to design of the study, risk of bias, inconsistent results, indirectness of evidence, imprecision, publication bias, the magnitude of the effect, existence of dose-response gradient, and if all plausible confoundings would only reduce the demonstrated effect.

## DISCUSSION

This systematic review and network meta-analysis aim to evaluate whether available evidence is enough to justify the use of some laryngoscopes (VLs or DLs) over the others for children’s orotracheal intubation. Current evidence in adults suggests VL to outperform DL for tracheal intubation and different VLs designs to have differential performance.^6^ However, such VL outperformance was not confirmed by previous meta-analyses of pediatric studies and there is little evidence regarding the relative performance of different laryngoscopes at present.^7,8^ We will then attempt to rank the diverse VLs and DLs by network meta-analyses trying to understand which devices perform better than others.

## Data Availability

Not applicable

## Data Availability

Not applicable

